# VULNERABLE CAROTID PLAQUE: ROBUSTNESS AND CLASSIFICATION CAPABILITIES OF MRI RADIOMIC FEATURES

**DOI:** 10.1101/2023.06.19.23291556

**Authors:** Zakaria Meddings, Leonardo Rundo, Umar Sadat, Xihai Zhao, Zhongzhao Teng, Martin J. Graves

## Abstract

**Objectives:** To assess how radiomic features may be combined with plaque morphological and compositional features identified by multi-contrast magnetic resonance imaging (MRI) to improve upon conventional risk assessment models in determining culprit lesions.

**Methods:** Fifty-five patients (mean age: 62.6; 35 males) with bilateral carotid stenosis who experienced transient ischaemic attack (TIA) or stroke were included from the CARE-II multi-centre carotid imaging trial (ClinicalTrials.gov Identifier: NCT02017756). They underwent MRI within 2 weeks of the event. Classification capability in distinguishing culprit lesions was assessed by machine learning. Repeatability and reproducibility of the results were investigated by assessing the robustness of the radiomic features.

**Results:** Radiomics combined with a relatively conventional plaque morphological and compositional metric-based model provided incremental value over a conventional model alone [area under curve (AUC), 0.819 ± 0.002 vs. 0.689 ± 0.019 respectively, p = 0.014]. The radiomic model alone also provided value over the conventional model [AUC, 0.805 ± 0.003 vs. 0.689 ± 0.019 respectively, p = 0.031]. T2-weighted imaging-based radiomic features had consistently higher robustness and classification capabilities compared with T1-weighted images. Higher-dimensional radiomic features outperformed first-order features. Grey Level Co-occurrence Matrix (GLCM), Grey Level Dependence Matrix (GLDM) and Grey Level Size Zone Matrix (GLSZM) sub-types were particularly useful in identifying textures which could detect vulnerable lesions.

**Conclusions:** The combination of MRI-based radiomic features and lesion morphological and compositional parameters provided added value to the reference-standard risk assessment for carotid atherosclerosis. This may improve future risk stratification for individuals at risk of major adverse ischemic cerebrovascular events.

**Clinical Relevance:** The clinical relevance of this work is that it addresses the need for a more comprehensive method of risk assessment for patients at risk of ischemic stroke, beyond conventional stenosis measurement. Radiomics provides a non-invasive means of assessing plaque vulnerability.

**Key points:**

- T2-weighted imaging-based radiomic features had consistently higher robustness and classification capabilities compared with T1-weighted images.
- Higher dimensional radiomic features had better performance than first-order features in identifying textures which could detect vulnerable carotid lesions.
- Radiomic features combined with MRI plaque features may improve atherosclerotic plaque risk stratification.

## INTRODUCTION

Carotid atherosclerotic disease is a subset of cardiovascular diseases (CVD), which is the leading cause of stroke and death worldwide [1]. Measuring the degree of luminal stenosis remains the major approach for clinical assessment of the severity of carotid atherosclerosis. It is the only currently validated diagnostic criterion for patient risk stratification. However, the majority of clinical events occur in patients with mild to moderate carotid stenosis [2]. Moreover, while the severe stenosis patients (≥70%) have been proven through clinical trials to benefit from carotid endarterectomy (CEA) and carotid artery stenting (CAS), the benefits of surgery taper off when stenosis severity is reduced (<70%) [2]. Therefore, those who lie in the mild-to-moderate stenosis range in particular require improved risk stratification for future ischaemic events.

Previous studies have suggested that morphological and compositional characteristics of atherosclerotic plaques may potentially better define clinical progression than luminal stenosis alone. A vulnerable carotid atherosclerotic plaque is often characterised by the presence of intra-plaque hemorrhage (IPH) and a large lipid-rich necrotic core (LRNC) with a thin or defective fibrous cap (FC) that can be visualized by high-resolution, multi-contrast magnetic resonance imaging (MRI) [3]. A comprehensive picture of plaque vulnerability should therefore include lesion morphological and compositional features [4,5]. Instead of simply identifying features which relate to atherosclerotic composition, radiomics may have the potential to add incremental value by using the data directly from images to identify features which describe lesions in a way which is beyond the reach of already established methods [6]. Some radiomic features may have obvious clinical relevance, such as the size of different zones within the image or the maximum brightness of an image; others are more abstract in nature, and represent patterns typically hidden to the human eyes.

In this study, it is hypothesised that radiomic features extracted from MRI can improve risk stratification of carotid plaques beyond conventionally used features such as the reference-standard degree of stenosis.

## MATERIALS AND METHODS

### Patients and image segmentation

Data from fifty-five patients with bilateral carotid stenosis from the Chinese atherosclerotic risk evaluation (CARE-II) multicentre clinical trial were used in this study (ClinicalTrials.gov Identifier: NCT02017756). IRB approvals were obtained for the entire study and for each participating institution, all participants in the study provided written informed consent [7]. The primary objective of CARE-II study was to determine the prevalence and characteristics of high-risk atherosclerotic plaques in the carotid arteries in Chinese patients with recent ischaemic stroke or transient ischaemic attack (TIA). In this study, linear logistic regression models were trained to differentiate culprit and non-culprit carotid artery lesions. The term ‘culprit’ always refers to the causative ipsilateral carotid artery or plaque, as determined following the assessment of ultrasound and MRI examinations. The ‘non-culprit’ side was deemed to have no causative basis for symptoms.

Those recruited underwent MRI of bilateral carotid arteries with the following sequences: three-dimensional (3D) time-of-flight (TOF), T1-weighted (T1) quadruple inversion recovery (QIR), T2-weighted (T2) multi-slice double inversion recovery (MDIR), and Magnetisation Prepared Gradient Recalled Echo (MPRAGE). Detailed MR imaging parameters can be found in the references [7], and in *Supplemental Material* A. Detailed patient demographics are provided in Table 1 below.

**Table 1.** Patient demographics (n=55).

|  |  |
| --- | --- |
| **Age, year (Mean ± SD)** | **62.6 ± 10.6** |
| **Smoker, n(%)** | 26 (47.3%) |
| **Male, n(%)** | 35 (63.6%) |
| **Hypertension, n(%)** | 36 (65.5%) |
| **Hyperlipidemia, n(%)** | 36 (65.5%) |
| **BMI, kg/m<sup>2</sup> (Mean ± SD)** | 24.9 ± 3.4 |

The MRI segmentations were performed manually by experts with reference to 3D time-of-flight (TOF), T1-weighted, T2-weighted and magnetisation-prepared rapid gradient echo (MP-RAGE) images to discern lumen and outer wall boundaries and various atherosclerotic components including IPH, LRNC and calcification [8]. The segmentation and related pre-processing procedures were as follows: (1) lumen and outer wall boundaries were drawn manually on the T1-weighted images to define the region of interest (ROI); (2) all images including the masks for the lumen and outer wall were converted into NIFTI format; (3) radio frequency bias correction [9] and de-noising using a spatially adaptive method [10] were performed; (4) global rigid and local deformable co-registrations were performed with T1-weighted images being the reference modality [11]; and (5) pixels of all images were finally extracted according to the segmentation masks. Except for Step (1), all other steps were performed automatically in MR-VascularView© (Nanjing Jingsan Medical Science and Technology, Ltd., Jiangsu, China).

### Radiomic feature extraction and robustness assessment

The PyRadiomics tool (v2.2.0) [12] was used to extract radiomic features based on the MRI pixels from slices which contained atherosclerotic disease defined by local plaque burden ≥ 50%. The feature families obtained by running multi-slice radiomic analysis were: shape features; first-order intensity histogram statistics; Grey Level Co-occurrence Matrix features (GLCM) [13–15]; Grey Level Run Length Matrix (GLRLM) [16]; Grey Level Size Zone Matrix (GLSZM) [17]; Grey Level Dependence Matrix (GLDM) [18]; and Neighbouring Grey Tone Difference Matrix (NGTDM) [19]. The default PyRadiomics setting, a fixed bin width of 25, was used. No resampling was applied.

The overall workflow is illustrated in Figure 1 and includes: calibration; pre-processing procedure to select robust features; development of the predictive model for the identification of culprit lesions; a post-processing procedure to select the most useful features; and the validation methodology. The dependence of radiomic features on the ROI, which defines the area from which features are extracted, was tested by measuring feature robustness using the intra-class correlation co-efficient (ICC) [20–22]. The ROI was manipulated by expanding and eroding the wall boundary outwards and inwards with 1-2 pixels. Only stable radiomic features would then be carried forward to the final analysis and those highly sensitive to minor ROI changes and image quality were omitted.

**Figure 1.**
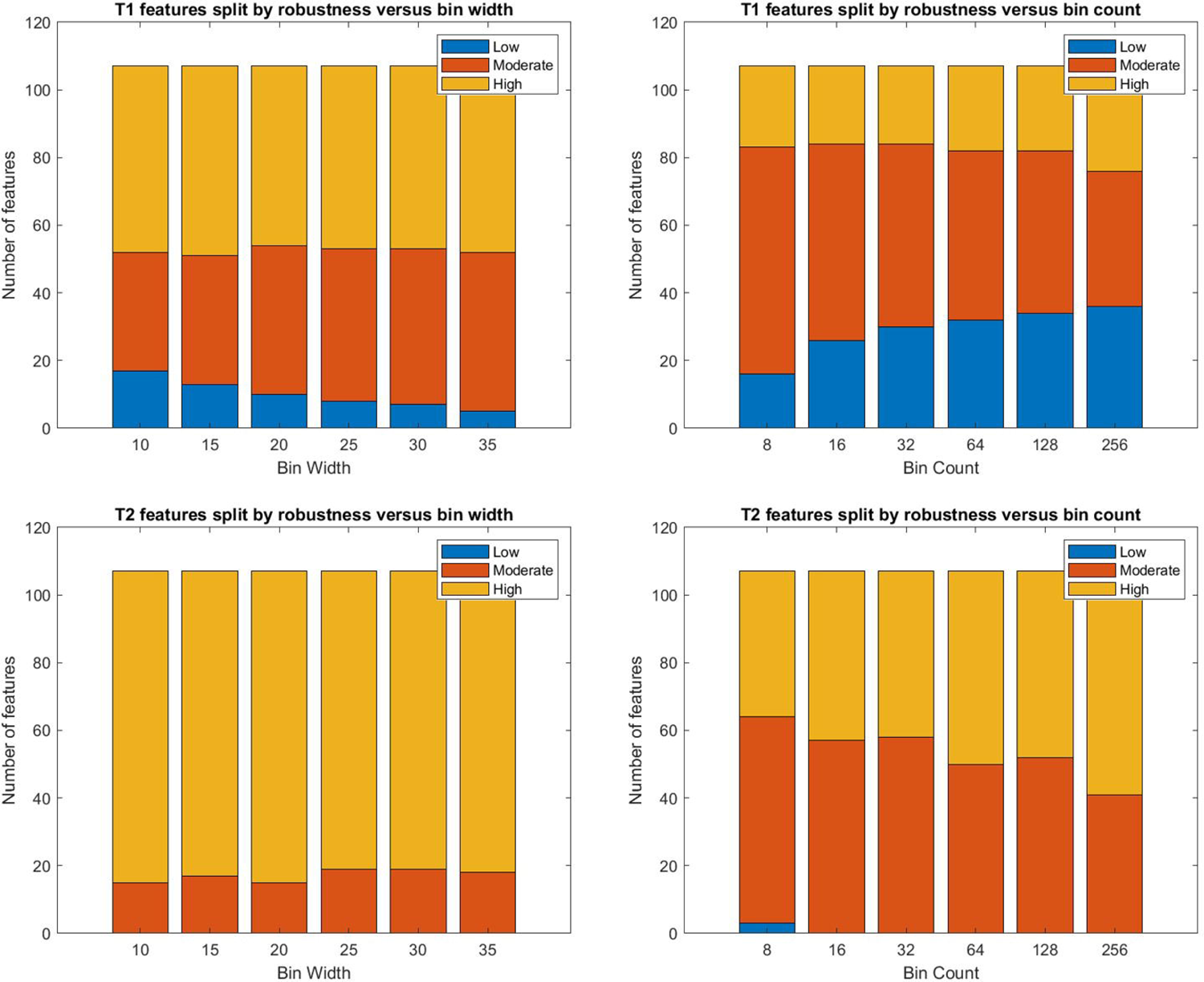
Schematic which demonstrates the methodology of extracting and evaluating radiomic features, and the cross-validation technique.

### Feature regularisation and model cross-validation

Least absolute shrinkage and selection operator (LASSO) and ridge regression were used to reduce over-fitting by introducing regularisation penalty terms to the cost function. In both LASSO and ridge regression, large co-efficients in the linear model are penalised by a characteristic penalty term, though in slightly different ways. LASSO regression involves the L1 norm of the vector of feature co-efficients w in its penalty term, which is the sum of the absolute values of the feature co-efficients. In ridge regression, the penalty term involves the square of the L2 norm, which is the sum of squares of feature co-efficients [23]. The corresponding penalty terms are shown below, where N refers to the number of features involved in the model.

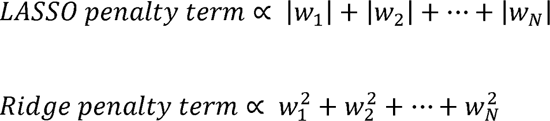

Since the sum of squares of co-efficients less than one is less than their sum, the ridge regression penalty is more accommodating for a more balanced co-efficient vector, while LASSO regression tends to favour multiple co-efficients being set to zero, resulting in a sparser model. While both types of regularisation were performed, with feature selection in mind, the results following LASSO regularisation were typically considered in the final analysis.

To validate the predictive ability of the model and assess its sensitivity to the dataset, a five-fold cross-validation scheme was employed where the data were split into five equally sized folds. Four fifths of the data are used to train a linear logistic regression model, and one fifth of the data are left unseen to evaluate the model. Additionally, to assess the repeatability of this analysis, this scheme was performed on resampled datasets 1,000 times according to the bootstrapping technique of resampling with replacement. Bootstrapping resampling measures the sensitivity to the data set of the cross-validation analysis. This also allowed for a more comprehensive analysis for feature selection.

### The combined model and feature selection

The combined model consisted of the radiomic model and conventional risk features composed of morphological and compositional metrics. Morphological metrics included: degree of stenosis; minimum minor axis length (MMAL); minimum lumen area (MLA); inward remodelling index (IRI); and outward remodelling index (ORI), plaque burden, and plaque volume. Compositional features included calcification presence, calcification volume, IPH presence, IPH volume, and LRNC presence, and LRNC volume. The merit of these features alone was considered in the univariate analysis.

To select features, a protocol was developed based on the cross-validation procedure used to identify the most useful features for the combined model. Using LASSO regularisation, the frequency of non-zero feature co-efficients across all five cross-validation folds and all bootstrap iterations was plotted to illustrate the features that were most commonly included across all possible models. With ridge regression, a similar principle was utilised, except that the features were considered selected if their corresponding average co-efficient across all five cross-validation folds was found to be in the top ten among all features considered in the model.

### Statistical analysis

Univariate analysis was performed using the χ^2^ test, Fisher’s exact test, or a student *t*-test when appropriate and those with p-value ≤ 0.1 were included in the multivariate analysis. The ICC was defined using a 2-way mixed analysis of variance (ANOVA) model. The relationship between sensitivity and specificity forms the basis of receiver-operating characteristic (ROC) curves. For all possible cut-offs (the point which separates culprit and non-culprit predictions) of the logistic regression S-curve, a point was added to the ROC curve and the resulting area under the curve (AUC) calculated accordingly. The AUC value is then used to measure the effectiveness of a given predictive model. The significance of the difference between two ROC AUC measurements were assessed using the Wilcoxon statistic for paired samples [24].

## RESULTS

### Univariate baseline predictors

The baseline model is firstly built upon the consideration of the univariate predictive value of morphological and compositional features. Geometric features except for ORI all had significant univariate predictive value, as did calcification presence and IPH presence. This is presented in Table 2 below. The multivariate predictive value of these features and the significance of the multivariate odds ratio within the model are also presented in Table 2.

**Table 2.** Univariate and multivariate analysis of the baseline model, consisting of lesion geometry and plaque morphology (n=55).

| Lesion geometry features | Univariate p-value | Multivariate Odds Ratio (95% CI) | Multivariate p-value |
| --- | --- | --- | --- |
| **Stenosis** | **0.009<sup>t</sup>** | 0.596 (0.161, 2.2) | 0.421 |
| **MMAL** | **0.0034<sup>t</sup>** | 1.708 (0.08, 34.2) | 0.717 |
| **MLA** | **0.036<sup>t</sup>** | 0.242 (0.014, 4.07) | 0.308 |
| **IRI** | **0.08<sup>t</sup>** | 2.315 (0.79, 6.73) | 0.111 |
| **ORI** | 0.735<sup>t</sup> | / | / |
| **Plaque Burden** | 0.102<sup>t</sup> | 1.213 (0.316, 4.659) | 0.771 |
| **Plaque Volume** | 0.18<sup>t</sup> | / | / |
| **Calcification Presence** | **0.07<sup>f</sup>** | **2.779 (1.196, 6.459)** | **0.014** |
| **Calcification Volume** | 0.33<sup>t</sup> | / | / |
| **IPH Presence** | **0.06<sup>f</sup>** | 0.954 (0.345, 2.637) | 0.925 |
| **IPH Volume** | 0.138<sup>t</sup> | / | / |
| **LRNC Presence** | 0.535<sup>c</sup> | / | / |
| **LRNC Volume** | 0.32<sup>t</sup> | / | / |

MMAL = minimum minor axis length; MLA = minimum luminal area; IRI = inward remodelling index; ORI = outward remodelling index. IPH = intra-plaque hemorrhage; LRNC = lipid-rich necrotic core; f = Fisher's exact test; t = Student's t-test; c = $\chi^2$ test.

### Robustness of radiomic features

Robustness of the radiomic features across 12 extraction settings (bin width: 10, 15, 20, 25, 30, 35 and bin count: 8, 16, 32, 64, 128, 256) was calculated for all 107 radiomic features produced by PyRadiomics. The T2-weighted images provided a higher proportion of moderate (0.5 ≤ ICC < 0.9) and high (ICC ≥ 0.9) robust features in comparison with T1-weighted images. Of the features derived from T1-weighted images, 20% had low robustness (ICC < 0.5), compared with almost none which were derived from T2-weighted images. The bin width was a more reliable extraction setting compared to the bin count, which led to a high number of low robustness features. This is presented in Figure 2.

**Figure 2.**
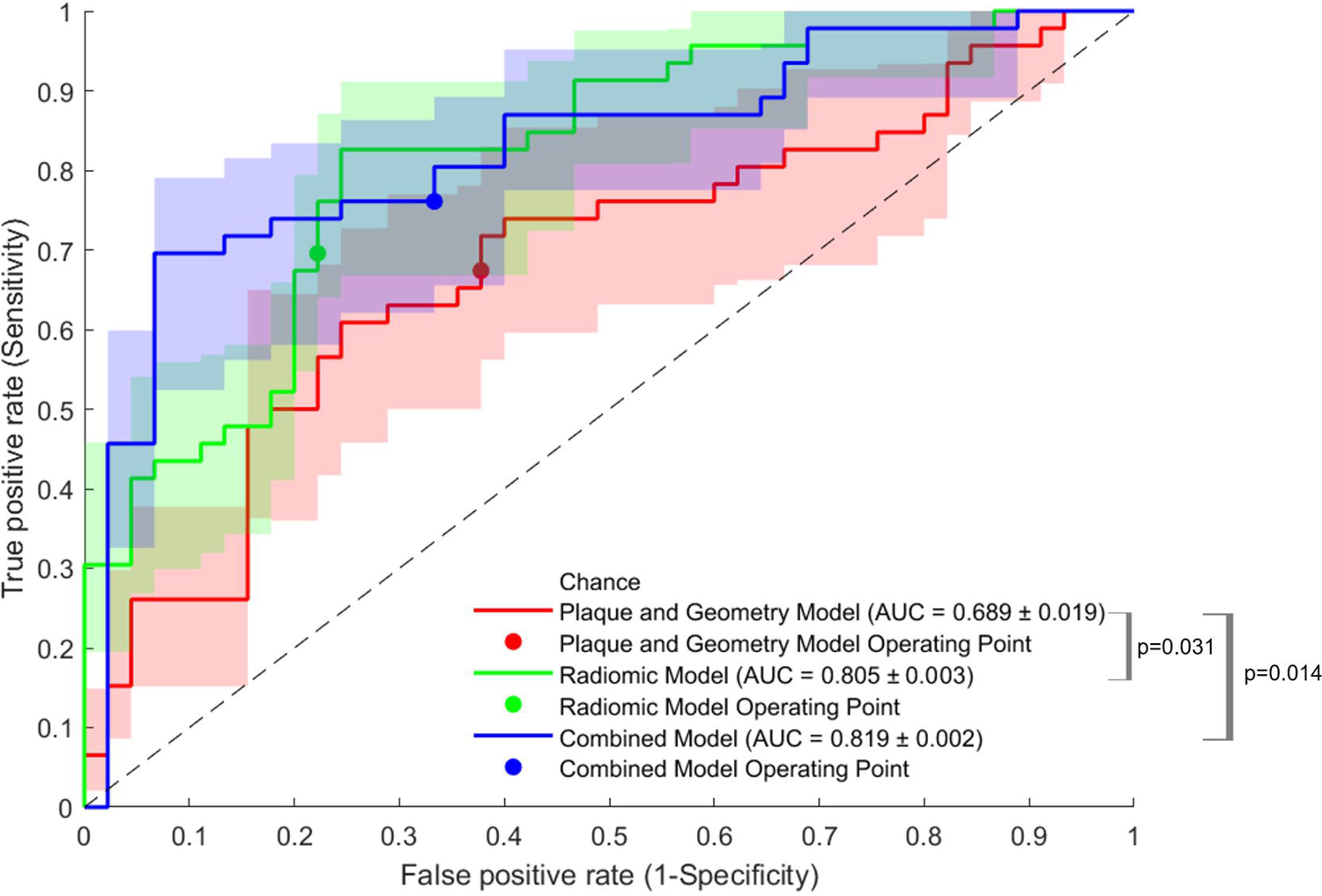
Robustness of T1- and T2-weighted images-derived radiomic features across 6 bin width and bin count settings (Low: intra-class correlation coefficient (ICC)<0.5; Moderate: 0.5≤ICC<0.9; High: ICC≥0.9).

### Integration of the radiomics and conventional models

The classification capabilities of models with different features included is tabulated in Table 3 across five cross-validation folds. It can be seen that if only lesion geometry (including stenosis) was considered, the overall accuracy was slightly greater than 60% at the optimal operating point, which is comparable to the case where only lesion components (calcification, etc) were considered.

**Table 3.** Classification capabilities of models with morphological and compositional features and radiomic features included and combined based on T2-weighted images.

| Classifier | AUC | Accuracy | PPV | NPV |
| --- | --- | --- | --- | --- |
| **Lesion geometry** | 0.638 ± 0.004 | 0.609 ± 0.010 | 0.610 ± 0.062 | 0.629 ± 0.022 |
| **Lesion composition** | 0.625 ± 0.004 | 0.644 ± 0.034 | 0.685 ± 0.042 | 0.646 ± 0.054 |
| **Geometry + composition** | 0.689 ± 0.019 | 0.684 ± 0.031 | 0.663 ± 0.042 | 0.729 ± 0.066 |
| **Radiomic model** | 0.805 ± 0.030 | 0.747 ± 0.030 | 0.745 ± 0.053 | 0.761 ± 0.050 |
| **Combined model** | 0.819 ± 0.020 | 0.758 ± 0.022 | 0.779 ± 0.064 | 0.754 ± 0.048 |
| **Simplified radiomic model** | 0.760 ± 0.011 | 0.712 ± 0.021 | 0.683 ± 0.021 | 0.753 ± 0.037 |
| **Simplified combined model** | 0.792 ± 0.015 | 0.778 ± 0.016 | 0.758 ± 0.009 | 0.802 ± 0.036 |

AUC = area under the curve; PPV = positive predictive value; NPV = negative predictive value.

The combination of lesion geometric and compositional features improved the classification capabilities, albeit without a significant difference when compared with either plaque morphology or composition alone. The radiomics model had better performance than any of the previous models. The combination of radiomic and lesion morphological and compositional features further improved the accuracy to 75.8%. The ROC plots relating to the performance of these models are illustrated in Figure 3.

**Figure 3.**
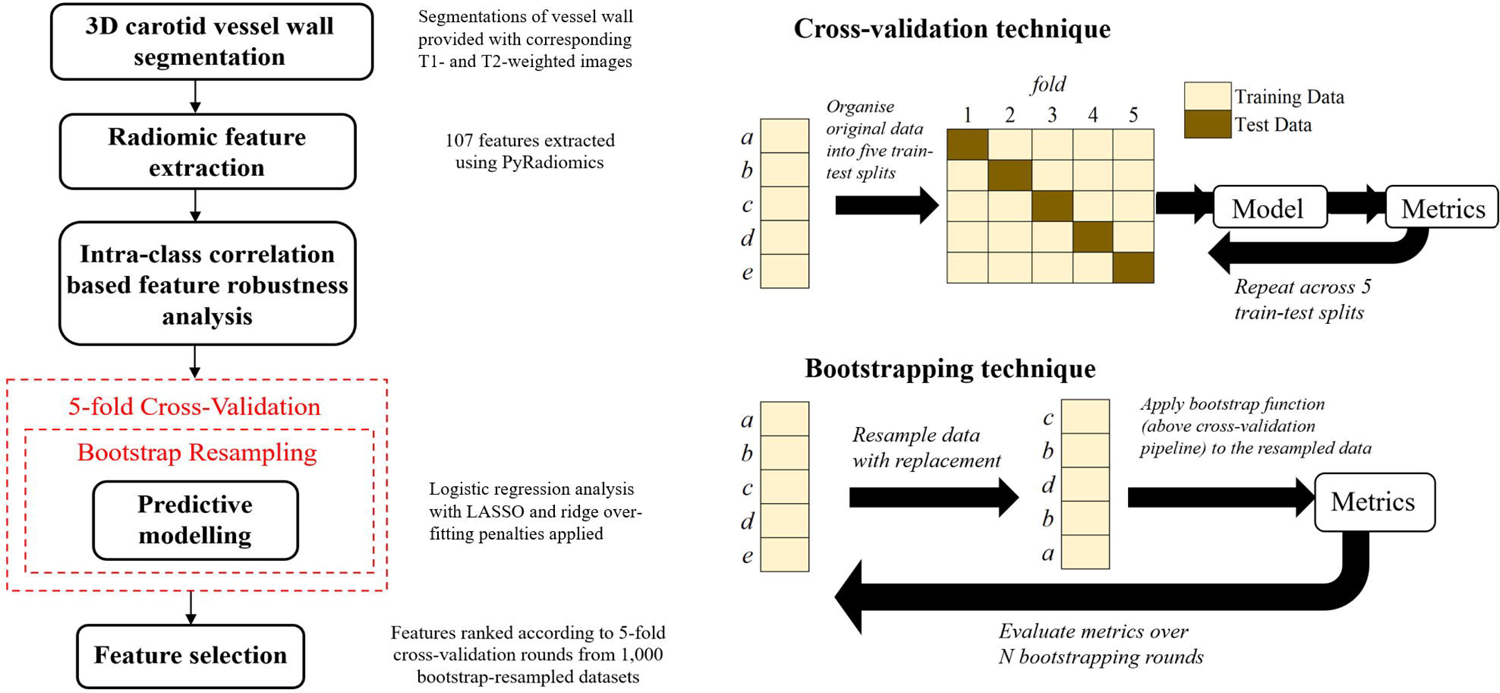
Receiver operating characteristic curves of the models considering lesion morphological and compositional features, and radiomic features alone and in combination.

The multivariate odds ratios of all features within the combined model are presented in *Supplemental Material* B, which highlights the relative weight of each feature within the cross-validated predictive models. This contains a conventional model, radiomic model, and a combined model.

In light of prioritising clinical interpretability, a simplified radiomic model was tested, which includes only seven radiomic features: Gray Level Non-Uniformity (GLSZM), Large Dependence High Gray Level Emphasis (GLDM), Small Dependence Low Gray Level Emphasis (GLDM), Difference Variance (GLCM), Informational Measure of Correlation (GLCM), kurtosis (first order), and skewness (first order). The performance of these simplified features is tabulated in Table 4 and the ROC plots may be found within *Supplemental Material* C, which contrasts the ability of simplified and full models (containing all available features) to distinguish culprit and non-culprit plaques.

**Table 4.**
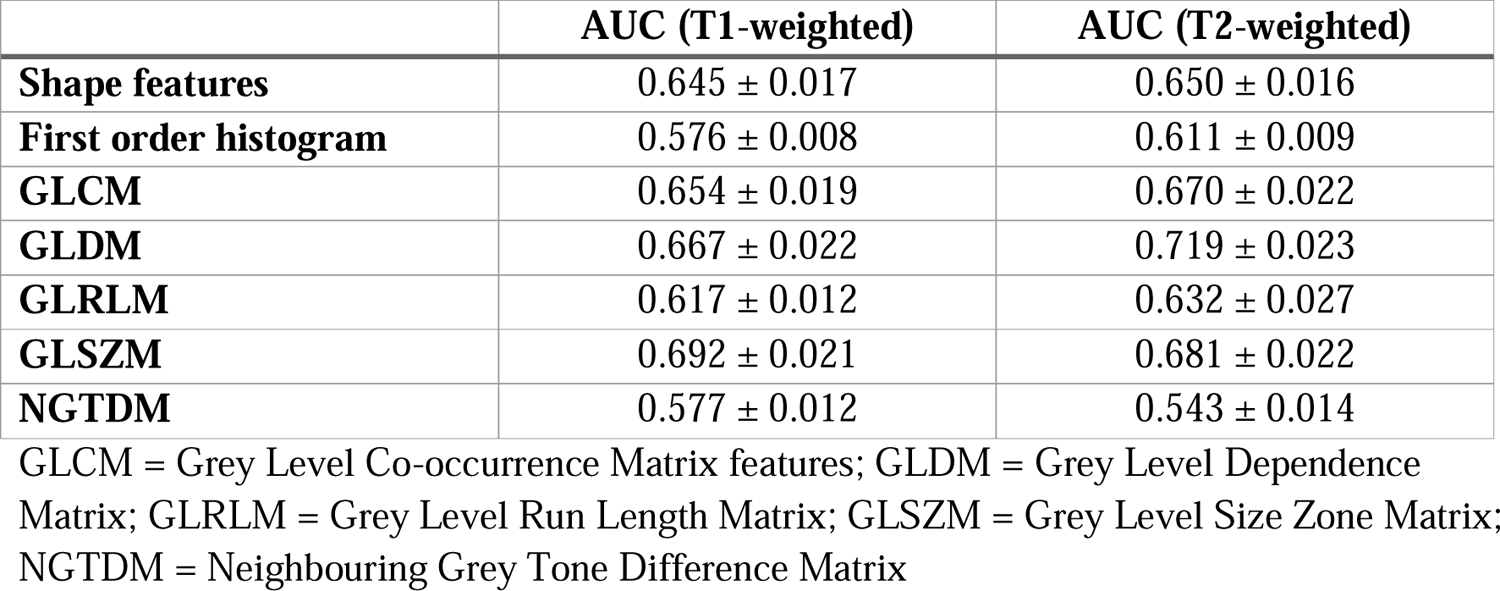
Classification capabilities of each family of radiomic features with cross-validation.

### Feature selection

Each feature class was analysed individually, and the corresponding ROC curves were created for calculation of the AUC. This is shown in Table 4. The AUC was calculated based on the performance of predictive models on a hold-out set according to the cross-fold validation method. In general, GLDM features derived from T2-weighted images had the best performance, followed by GLSZM and GLCM. NGTDM features had the poorest predictive value, and the poorest robustness profile.

The frequency of selection using LASSO and ridge protocols developed in this study are presented in the *Supplemental Materials* D and E. First order features skewness and kurtosis appear consistently in feature selection according to the established protocol, despite the first-order feature family in itself not having the strongest predictive performance. The morphological and compositional features including calcification, IPH and plaque burden (PB) were often selected by the linear model under different perturbations of the training data.

## DISCUSSION

While univariate analysis revealed the capability of stenosis and other more intricate geometrical features to differentiate culprit and non-culprit plaques, these assessments do not tell the whole story. Features obtained using MRI improved upon the predictive capability of stenosis measurement alone. Radiomics then further improved the predictive model to provide a robust means of describing the plaque vulnerability. This was also true of a simplified model with the involvement of GLSZM, GLDM and GLCM features, kurtosis, and skewness. This implies that interpretable features become more useful when combined with other higher order features along with the plaque features; it also emphasises the multifactorial, complex nature of carotid plaque vulnerability. T2-weighted imaging-based radiomic features had consistently higher robustness than T1-weighted images as measured by the ICC among radiomic features. This may have then carried through to the superior classification capabilities of features derived from the T2-weighted images as opposed to the T1-weighted images. This suggests that T2-weighting may be more efficient at picking up tissue contrasts which relate to plaque vulnerability.

Since the plaque components such as IPH and the extracellular fibrous matrix can take multiple forms, it is possible that radiomic texture features detect the particular sub-types of these plaque features which are more vulnerable. The profile of plaque haemorrhage - for instance, how it has developed and progressed - as identified by its texture features, may be a more powerful tool to investigate plaque vulnerability than knowledge of its presence alone. The high recurrence of inclusion of IPH and PB in the linear models following bootstrap resampling re-enforces the notion that the application of AI in carotid plaque vulnerability detection should be seen as a tool to incrementally improve risk stratification, since the radiomic features provided complementary information to the traditional plaque-based model – the features in the plaque morphological and compositional model ultimately remain more clinically interpretable than even simpler radiomic features, since they relate directly to pathology.

The model which combined the full radiomics signature with plaque morphological and compositional information provided a significant improvement in AUC [AUC, 0.819 ± 0.002 vs. 0.689 ± 0.019 respectively, p = 0.014], while the radiomic model alone also provided significant improvement over the conventional model [AUC, 0.805 ± 0.003 vs. 0.689 ± 0.019 respectively, p = 0.031]. On closer inspection, though the discrepancy in the AUC increase between the radiomic-only and all-inclusive models is relatively low, with a mean AUC increase of 1.74%, the statistical significance was improved by 98.6%, which increases the certainty of improvement over conventional methods when radiomic features are combined with the morphological and compositional features.

Limitations include the retrospective nature of the analysis and number of patients. Imaging was performed post-event, and the analysis aims to formulate a means of predicting patient outcome before the event occurs. Classifiers may not be so meaningful if the rupture has already happened and caused significant remodelling within the timeframe between event and patient presentation. Therefore, this work relies upon the implicit assumption that the underlying morphology is highly similar to what it was in the time leading up to the event. In the CARE-II trial, the MRI protocol was pre-defined. The 2mm slice thickness could incur information loss. The effect of different image quantisation settings was explored, but how well the radiomic features would perform under MRI scanners with different signal-to-noise ratio and other imaging parameters remains to be seen. Image pixel intensity normalisation during pre-processing was forwent due to homogeneous image acquisition parameters. External validation on different scanners would be required to validate the robustness of the risk prediction models recommended by this study. Different ROI delineations, as defined by the original manual segmentation, impact the radiomic feature extraction. Perturbations of the original ROI using dilation and erosion allows us to understand which image modalities are robust to inter-observer variability. Radiomic features extracted from T2-weighted images were generally more robust to these ROI perturbations.

In conclusion, risk stratification for carotid atherosclerosis can be improved by a more comprehensive overview of the plaque risk beyond the current gold-standard degree of stenosis measurement. These results suggest that the geometric and plaque compositional features, in conjunction with radiomic features, can provide diagnostic improvement for patients with carotid atherosclerosis.

## Data Availability

All data produced in the present study are available upon reasonable request to the authors

## Acknowledgments

This research was supported by the NIHR Cambridge Biomedical Research Centre (BRC-1215-20014). The views expressed are those of the authors and not necessarily those of the NIHR or the Department of Health and Social Care. MJG receives funding from the NIHR Cambridge Biomedical Research Centre (BRC-1215-20014) and ZM receives funding from the Cambridge Trust (10468740).

## Abbreviations and acronyms

ANOVA: analysis of variance

AUC: area under the curve

CARE: Chinese atherosclerotic risk evaluation

CAS: carotid artery stenting

CEA: carotid endarterectomy

CVD: cardiovascular disease

FC: fibrous cap

GLCM: grey level co-occurrence matrix

GLRLM: grey level run length matrix

GLSZM: grey level size zone matrix

GLDM: grey level dependence matrix

NGTDM: neighbouring grey tone difference matrix

MLA: minimum lumen area

MMAL: minimum minor axis length

MP-RAGE: magnetisation prepared rapid gradient echo

MRI: multi-contrast magnetic resonance imaging

ICC: intra-class correlation co-efficient

IPH: intra-plaque haemorrhage

IRI: inward remodelling index

LASSO: least absolute shrinkage and selection operator

LRNC: lipid-rich necrotic core

MDIR: multi-slice double inversion recovery

MLA: minimum lumen area

MMAL: minimum minor axis length

NPV: negative predictive value

OR: odds ratio

ORI: outward remodelling index

PPV: positive predictive value

QIR: quadruple inversion recovery

ROC: receiver operating characteristic

ROI: region of interest

TIA: transient ischemic attack

TOF: time of flight

