## Supplemental Material for "VULNERABLE CAROTID PLAQUE: ROBUSTNESS AND CLASSIFICATION CAPABILITIES OF MRI RADIOMIC FEATURES"

**An evaluation of HR-MR radiomic features in identifying carotid plaque vulnerability – Supplementary Material**

**Supplementary Material A: High-Resolution carotid MRI acquisition details**

The recruited patients underwent multi-centre bilateral MRI of the carotid arteries using 3.0 T MRI scanners (Philips Achieva TX) each with an 8-channel phased-array carotid artery coil [7]. Uniformity of imaging was achieved using a multi-contrast vessel wall imaging protocol. This includes three-dimensional (3D) time-of-flight (TOF), T1-weighted (T1w) quadruple inversion recovery (QIR), T2-weighted (T2w) multi-slice double inversion recovery (MDIR), and Magnetisation Prepared Gradient Recalled Echo (MP-RAGE) imaging sequences. The localisation of carotid plaque imaging was centred to the bifurcation of index carotid artery.

The common MRI scanning parameters employed were: (1) 3D TOF: TR/TE=0.02/0.006 ms, field of view (FOV) = 150x150 mm^2^, matrix = 512x512, slice thickness = 2 mm, NEX=1, sequence duration = 5 minutes; (2) T1: TR/TE=800/10 ms, field of view (FOV) = 150x150 mm^2^, matrix = 512x512, slice thickness = 2 mm; (3) T2: TR/TE=4800/50 ms, field of view (FOV) = 150x150 mm^2^  , matrix = 512x512, slice thickness = 2 mm.

**Supplementary Material B: Multivariate Odds Ratio Table**

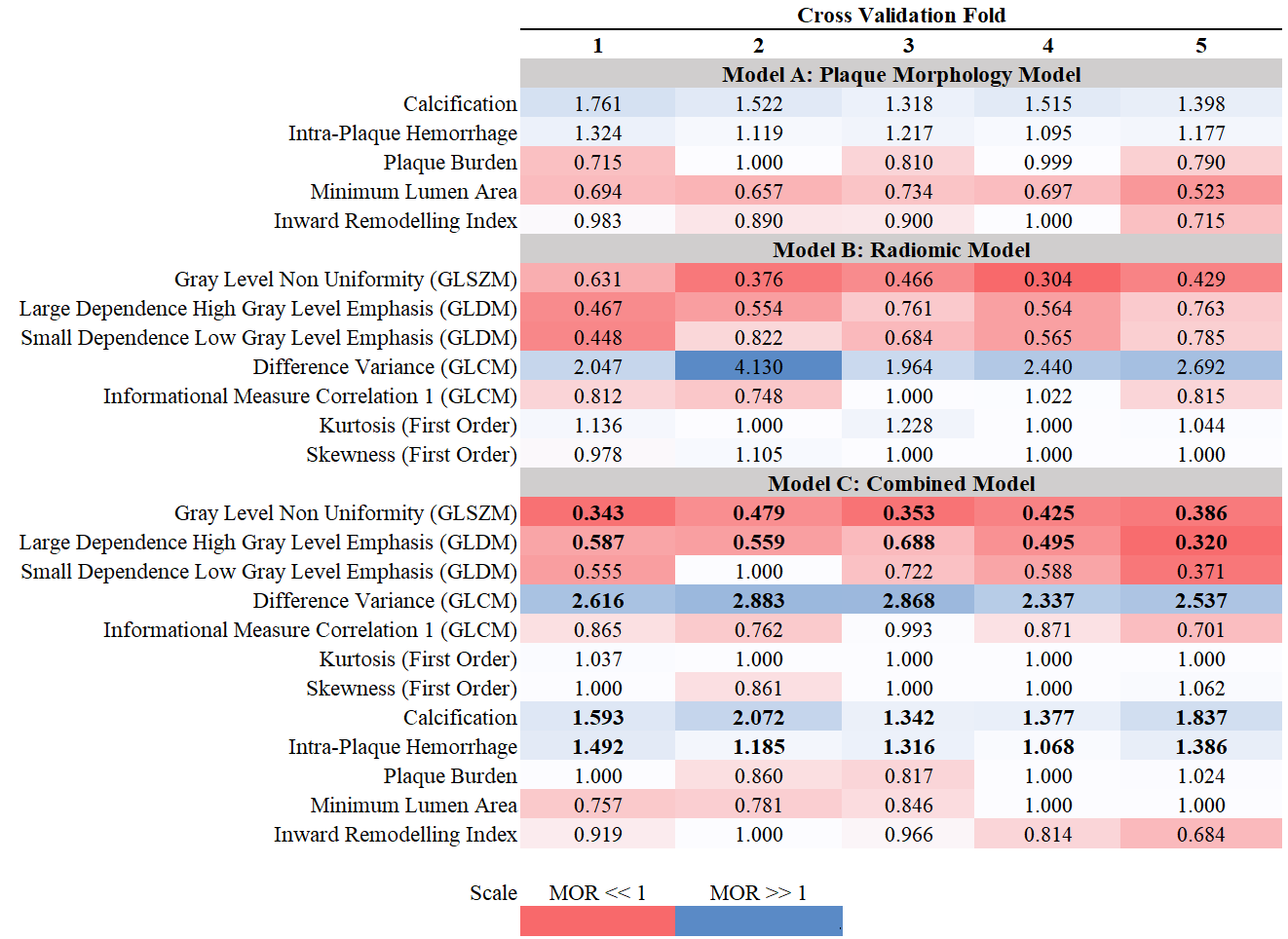

**Supplementary Material C: ROC Plots**

**Combined Model versus Conventional Model**

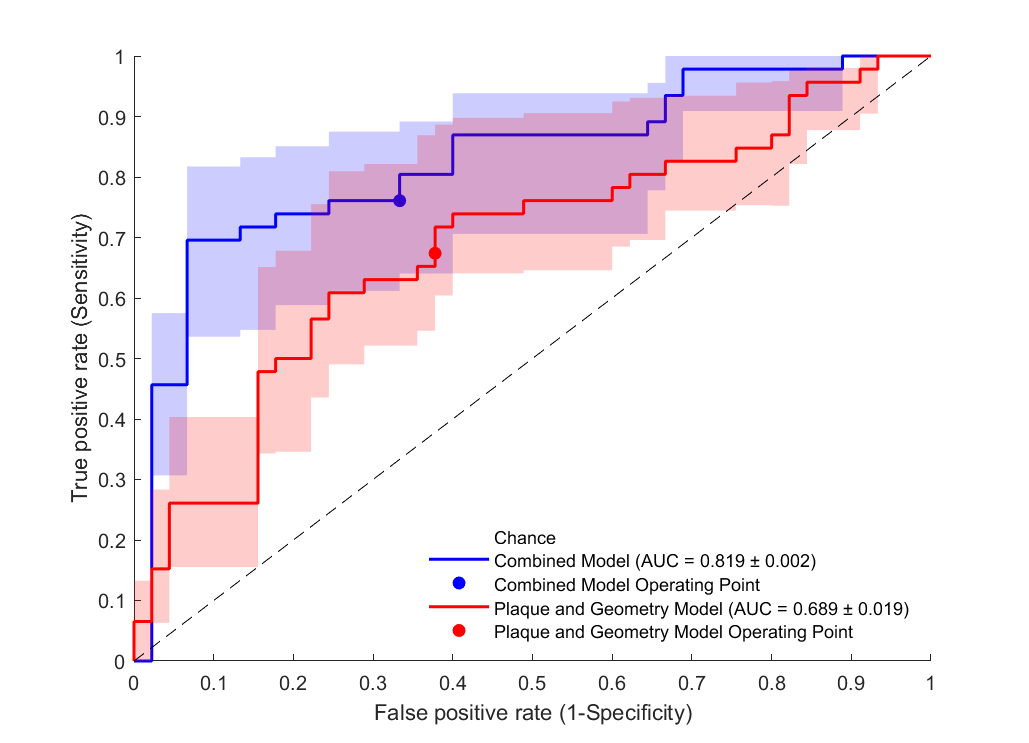

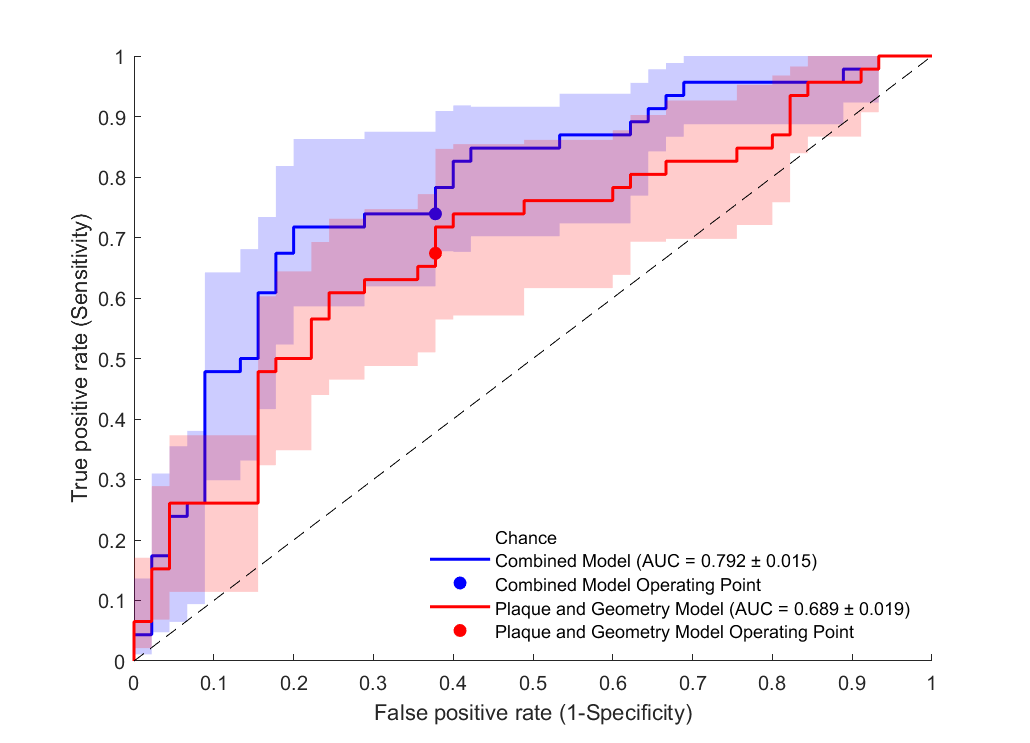

**Simplified Combined Model versus Conventional Model**

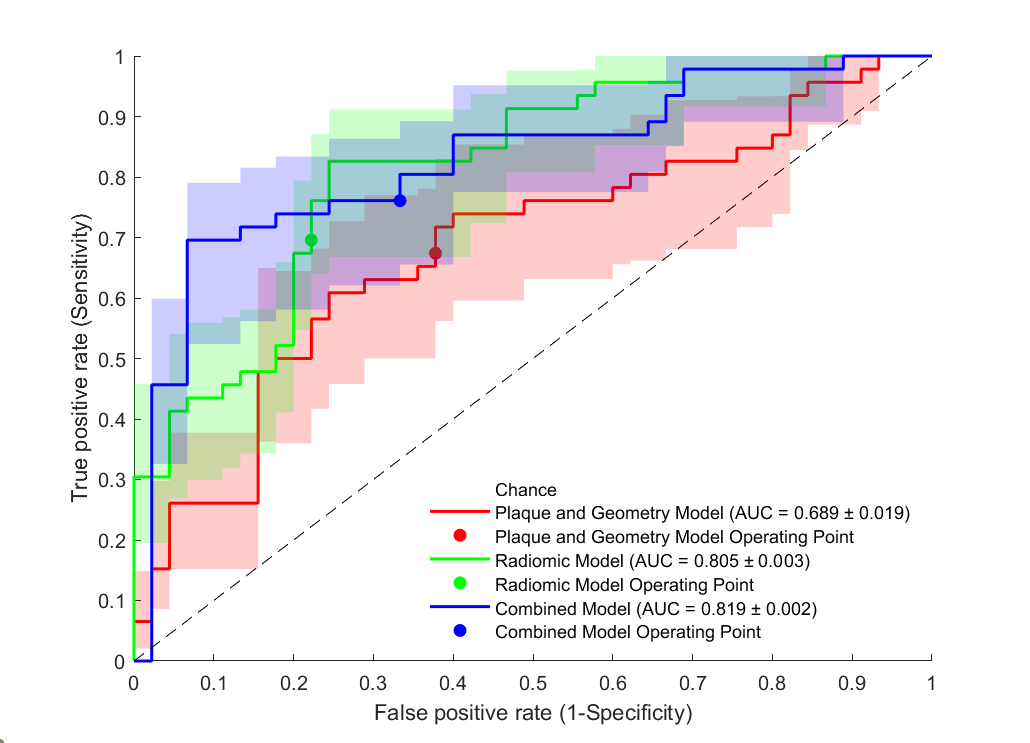

**Combined Model versus Radiomic and Conventional Models**

**Simplified Combined Model versus Radiomic and Conventional Models**

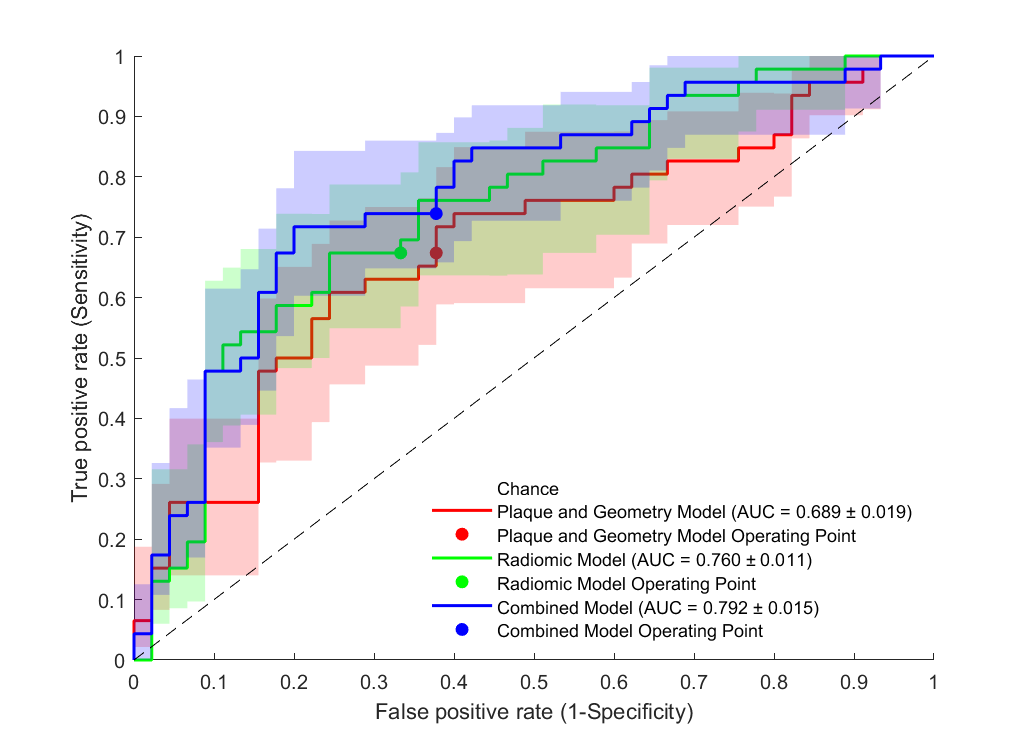

**Supplementary Material D: Selection Frequency - Ridge**

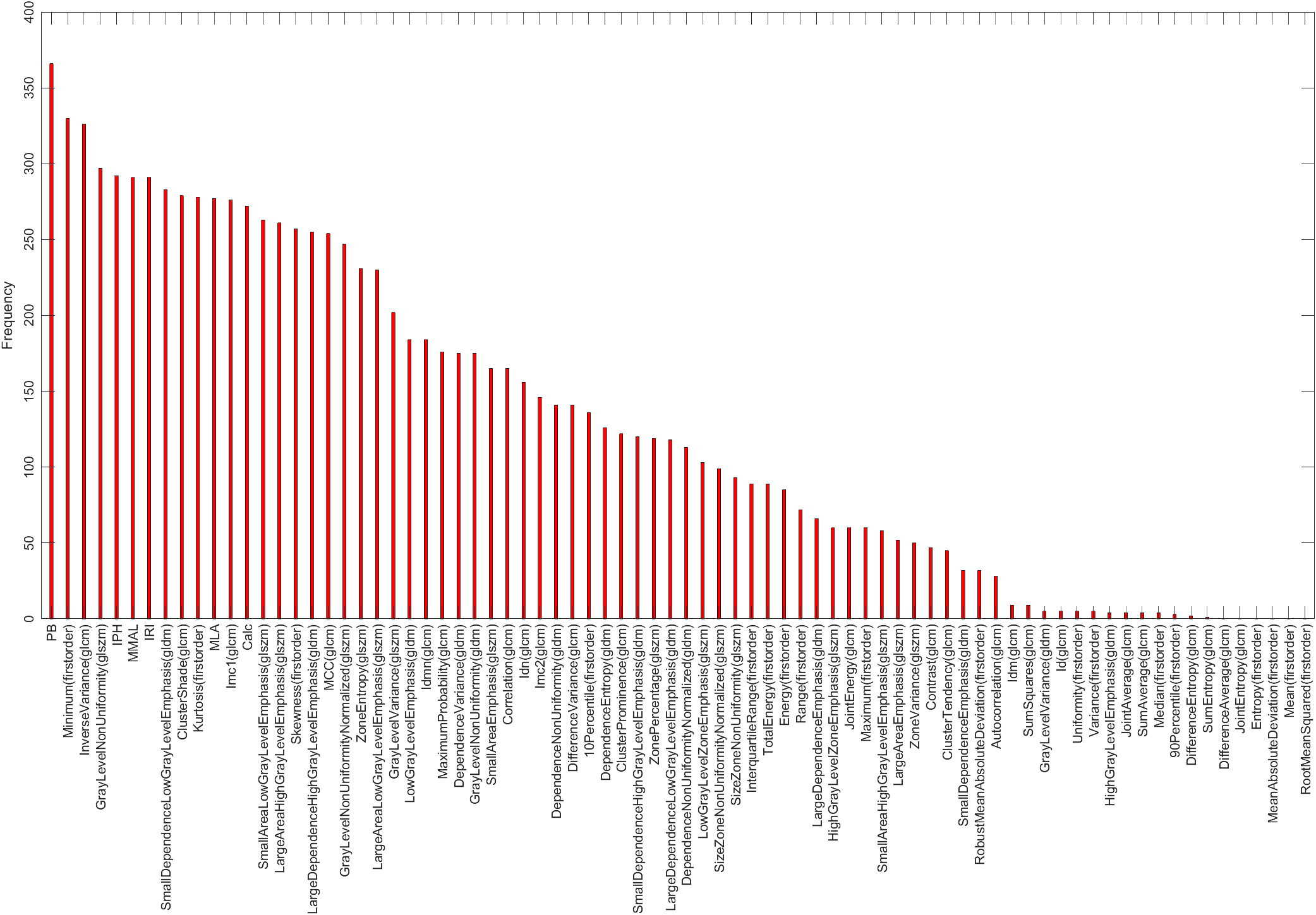

**Supplementary Material E: Selection Frequency – LASSO**

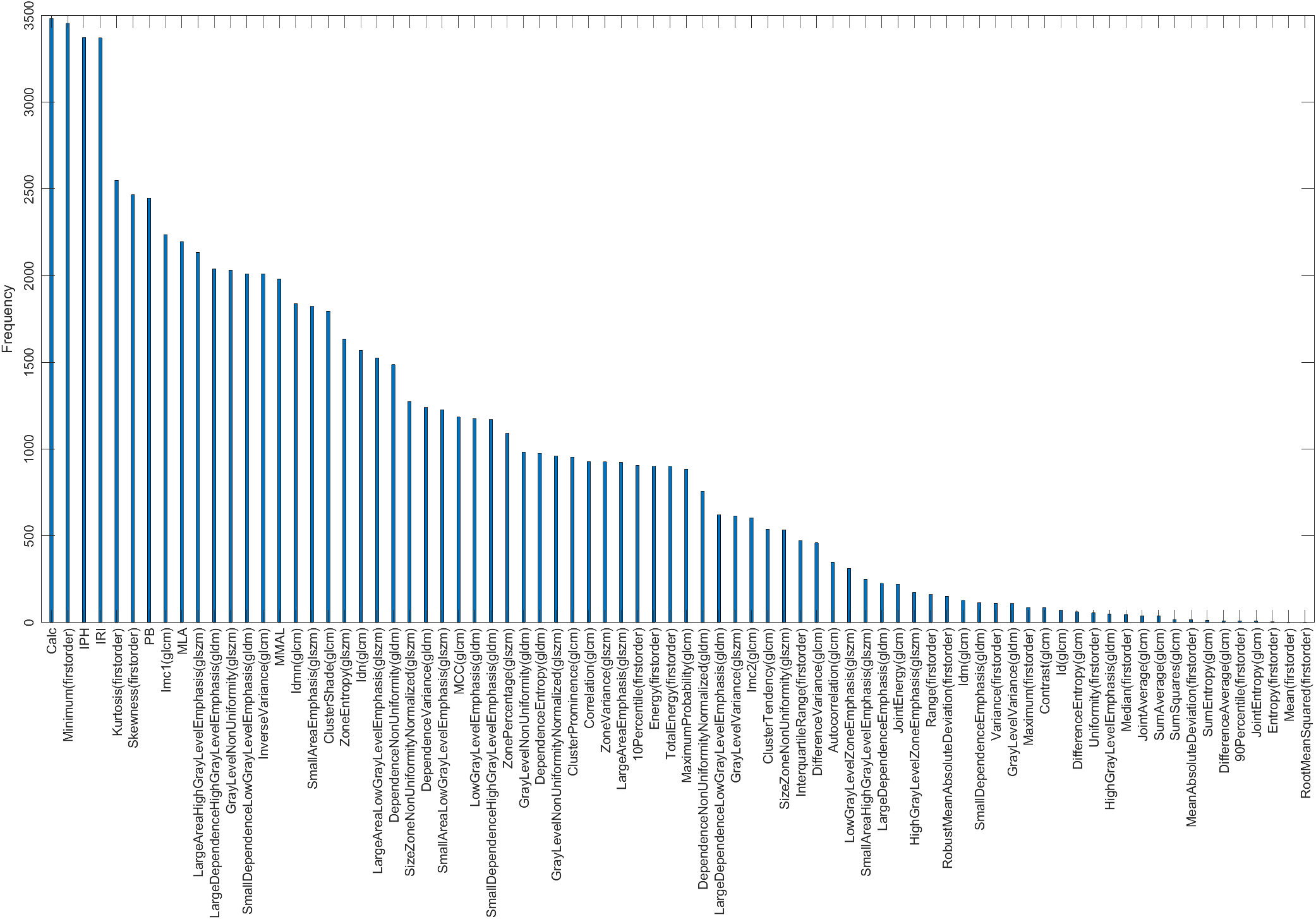
